# Plasma ptau217, NfL, GFAP diagnostic performance and biomarker profiles in Alzheimer disease, frontotemporal dementia, and psychiatric disorders, in a prospective unselected neuropsychiatry memory clinic

**DOI:** 10.1101/2025.07.09.25331224

**Authors:** Dhamidhu Eratne, Matthew Kang, Charles B Malpas, Christa Dang, Courtney Lewis, Oneil G Bhalala, Qiao-Xin Li, Steven Collins, Colin L Masters, Samantha M Loi, Alexander F Santillo, Kaj Blennow, Henrik Zetterberg, Dennis Velakoulis, The MiND Study Group

## Abstract

**INTRODUCTION:** Plasma biomarkers offer promise for improving diagnosis of Alzheimer’s disease (AD) and differentiating AD and other neurodegenerative disorders (ND) like frontotemporal dementia (FTD) from primary psychiatric disorders (PPD), particularly in younger patients.

**METHODS:** In this prospective study, we investigated plasma phosphorylated tau 217 (ptau217), neurofilament light chain (NfL), and glial fibrillary acidic protein (GFAP) in 341 unselected participants from a neuropsychiatry memory clinic, including AD (n=40), bvFTD (n=15), PPD (n=69), other ND, and controls.

**RESULTS:** Plasma ptau217 showed strong diagnostic performance for distinguishing AD from bvFTD (96% accuracy) and PPD (93% accuracy). NfL best distinguished all ND from PPD, while GFAP showed limited diagnostic utility. Biomarker profiles using predefined cut-offs and age-adjusted z-scores further clarified group differences.

**DISCUSSION:** Plasma ptau217 and NfL have strong diagnostic utility in real-world, diagnostically complex cohorts. These findings support implementation of scalable blood-based biomarkers to improve early and accurate diagnosis in memory clinic settings.

## INTRODUCTION

There has been a dramatic increase in potential of blood-based biomarkers to transform diagnosis and clinical care of patients presenting with cognitive, psychiatric, and neurological symptoms. Such potential includes improving early and accurate diagnosis of Alzheimer’s disease, and importantly, helping resolve the diagnostic difficulties in real-world clinical settings with diverse neuropsychiatric presentations, such as distinguishing Alzheimer disease (AD) from other causes of neurodegeneration such as behavioural variant frontotemporal dementia (bvFTD), and from common differential diagnostic dilemmas such as primary psychiatric disorders (PPD) [1–6]. Early, accurate diagnosis is even more important now in this era of increasing availability of disease specific treatments, such as monoclonal antibodies targeting beta amyloid. Two of the most promising biomarkers in this regard are plasma phosphorylated tau 217 (ptau217), and neurofilament light chain protein (NfL). Ptau217 has been shown in several studies to have strong diagnostic performance and specificity, to distinguish AD from non-AD disorders (such as non-AD dementias, and PPD) [7–10].

Within AD and control cohorts, plasma levels of ptau217 increase with more severe tau pathology assessed by tau PET [11] or tangle load at autopsy [12], and plasma ptau217 is increased in disorders with tangles but without amyloid plaques [13], supporting that it reflects tau phosphorylation state and tau pathology. As a non-specific marker of neurodegeneration of especially long myelinated axons in the CNS as well as acute neuronal injury, NfL has been shown to strongly distinguish neurodegenerative disorders (ND), from ND mimics including PPD and non-neurodegenerative disorders [3,4,14,15], and to have utility in clinical settings [14,16–19]. Glial fibrillary acidic protein (GFAP), a marker of astrocytic activation and neuroinflammation, is an additional biomarker of interest [14,20–26]. However, the utility of GFAP for the clinical differentiation between AD, non-AD ND, and PPD, especially when compared to ptau217 and NfL is not yet clear.

We have previously described the diagnostic performance, and a range of cut-offs and reference ranges for each of plasma ptau217 and NfL [10,14,18,27–29], but have not investigated their utility when combined. Furthermore, while we have published data showing the the superiority of NfL compared to glial fibrillary acidic protein (GFAP) to distinguish bvFTD from PPD [30], we have not yet investigated all three, ptau217, NfL, GFAP, in a younger real-world clinical neuropsychiatric cohort with diverse ND and PPD. A few studies have investigated combinations of these biomarkers and different biomarker profiles. Benussi et al investigated NfL, ptau217, and NfL/ptau217 ratios, and other biomarkers, in 374 participants (97 AD, 278 FTLD) [31]. They found strong diagnostic performance of ptau217 and slightly superior performance of NfL/ptau217 to distinguish between these two groups, although they but they lacked a PPD group, which is critical to appreciate use of biomarkers for bvFTD. Rousset et al investigated NfL and ptau217 in an unselected memory clinic setting, finding strong utility of ptau217 for AD diagnosis. They also described high/low ptau217 / high/low NfL profiles in AD and several clinical diagnostic groups [17]. To our knowledge, no studies to date have investigated ptau217, NfL, GFAP, and biomarker profiles, with a specific focus on AD, bvFTD, PPD, in younger people, where differential diagnose, diagnostic uncertainty, misdiagnosis, and delay, are greater [1,32–34]. Further research is needed to properly establish the roles of single and combination biomarkers in real-world clinical settings with diverse neuropsychiatric cohorts.

The primary aim of this study was to compare levels and diagnostic performance of plasma ptau217, NfL, ptau217:NfL ratio, and GFAP, for distinguishing between AD, bvFTD, and PPD. The hypothesis was that combination ptau217 and NfL would increase diagnostic distinction, compared to either biomarker alone. In addition we aimed to compare diagnostic performance of ptau217, NfL, GFAP to distinguish ND from PPD. Finally, we aimed to describe ptau217/NfL biomarker profiles of AD pathology and neuronal injury (e.g. low AD/low neuronal injury, high AD/high neuronal injury) in an unselected population of patients from a neuropsychiatry memory clinic. We focussed on ptau217 and NfL, using our previously described cut-off for plasma ptau217 [10], and age-adjusted z-score reference range models for plasma NfL [27,30]. While a few studies have published age-binned reference ranges for GFAP in serum [35] and plasma [36], we are not aware of any studies that have described GFAP reference ranges using similar modelling for precise age-adjusted percentiles and z-scores, like we did for NfL. As an exploratory aim, we also described ptau217/NfL/GFAP (AD/neuronal injury/neuroinflammation) profiles, by creating a novel age-adjusted z-score model for plasma GFAP across the lifespan.

## METHODS

The current study included participants prospectively recruited between June 2019 and April 2023, who had provided a blood sample for biomarker analysis. This is a follow up on our previous study cohort in which we investigated plasma and CSF NfL [14]. In the current study, we included participants who had plasma ptau217, NfL, and GFAP biomarker results available. In the current study the focus was on AD, bvFTD, PPD diagnostic groups, with comparator groups being control subjects and Other ND (both described in the previous study). Data was available in the current study for additional comparator groups: people with mild cognitive impairment, and people with presymptomatic genetic ND (not previously described).

Participants were recruited from the Neuropsychiatry Centre at The Royal Melbourne Hospital, a quaternary service receiving referrals for diagnostically complex cases from primary care and other specialist services within Australia. Participants, as part of routine clinical care through the Neuropsychiatry Centre, received comprehensive multidisciplinary assessments and multimodal investigations, including CSF AD biomarker analysis, with gold standard consensus diagnosis based on established diagnostic criteria, as previously described in detail [14,18,28]. Control participants were people recruited from the community, with no symptoms or diagnoses of neurological or neurodegenerative disorders, no active psychiatric symptoms or conditions. Diagnostic group categorisation was determined based on most recent diagnosis, at longitudinal follow up, based on established diagnostic criteria, blinded to plasma biomarker levels, as previously described [14,18,28].

EDTA plasma samples were collected during patients’ diagnostic workup, and at first visit for community controls, and samples were stored at –80C. Plasma NfL and GFAP were measured using N2PB kits on a Quanterix Single molecule array (Simoa) HD-X analyzer, according to the manufacturer’s instructions (Quanterix Corporation, Billerica, MA, USA). Plasma ptau217 was measured using an in-house University of Gothenburg (UGOT) ptau217 assay, as previously described in detail [37]. The measurements were performed in one round of experiments using one batch of reagents by analysts blinded to clinical data and diagnoses, thereby reducing potential batch effects.

This study, part of The Markers in Neuropsychiatric Disorders Study (The MiND Study, https://themindstudy.org), was approved by the Human Research Ethics Committee at Melbourne Health (2016.038, 2017.090, 2018.371, 2020.142).

### Statistical analyses

Statistical analyses were performed using R version 4.5.0 (2025-04-11). Biomarker levels in different groups were compared using standardised bootstrapped general linear models (GLM), with age at blood sample and sex as additional covariates. ROC curve analyses were then performed to investigate diagnostic utility between different combinations of groups. Bootstrapped differences in AUC were used to compare ROC curves. Optimal cut-offs were selected based on Youden’s *J* statistic. Additional diagnostic test parameters were computed: positive and negative likelihood ratios, positive and negative predictive values, overall accuracy, and diagnostic odds ratio. Additional sensitivity analyses were performed: excluding extreme outliers and performing all GLMs with weight included as a covariate. As results were similar, the results excluding weight were presented to maximise the sample sizes for analyses and presented results (since not all participants had weight data).

We focused on describing ptau217 and NfL biomarker profiles. These were Low ptau217/ Low NfL (a low AD pathology and low neuronal injury profile – a “normal” profile), and abnormal profiles: Low ptau217/High NfL (low AD pathology but elevated neuronal injury), High ptau217/Low NfL (AD pathology but low neuronal injury), and High ptau217/High NfL (high AD pathology and high neuronal injury). Plasma ptau217 and NfL biomarker levels were dichotomised in to “High” and “Low”, based on our previously published data and cut-offs that used the same assays [10,27]. For ptau217, we used the cutoff of 2.35 that was optimal at distinguishing AD from non-AD, as previously described and published [10]. For plasma NfL, given the strong non-linear association with age, we used age-based percentiles and z-scores derived from the generalised additive models for location, scale, and shape (GAMLSS) model of a large reference control cohort that we developed, previously described and published [14], defining the 95^th^ percentile as the cut-off between “High” and “Low”. For this study, we created a novel age-adjusted GFAP reference range model using (GAMLSS), using the control group from this study. This allowed us to derive more precise age-based percentiles and z-scores and more precise characterisation, again given the significant strong and non-linear association with age compared to coarse age-binned cut-offs. The 95^th^ percentile was defined as the cut-off for “High” and “Low” GFAP levels.

## RESULTS

The final cohort consisted of 341 participants: 40 with Alzheimer disease (median age 62 years, 53% female), 15 with bvFTD (median age 57 years, 27% female), 69 with PPD (median age 55 years, 48% female). There were 119 controls (median age 63, 74% female), and in the additional comparator groups: 67 with other ND, 13 with MCI, and 18 with presymptomatic genetic ND (see Table 1 for full details).

**Table 1.**
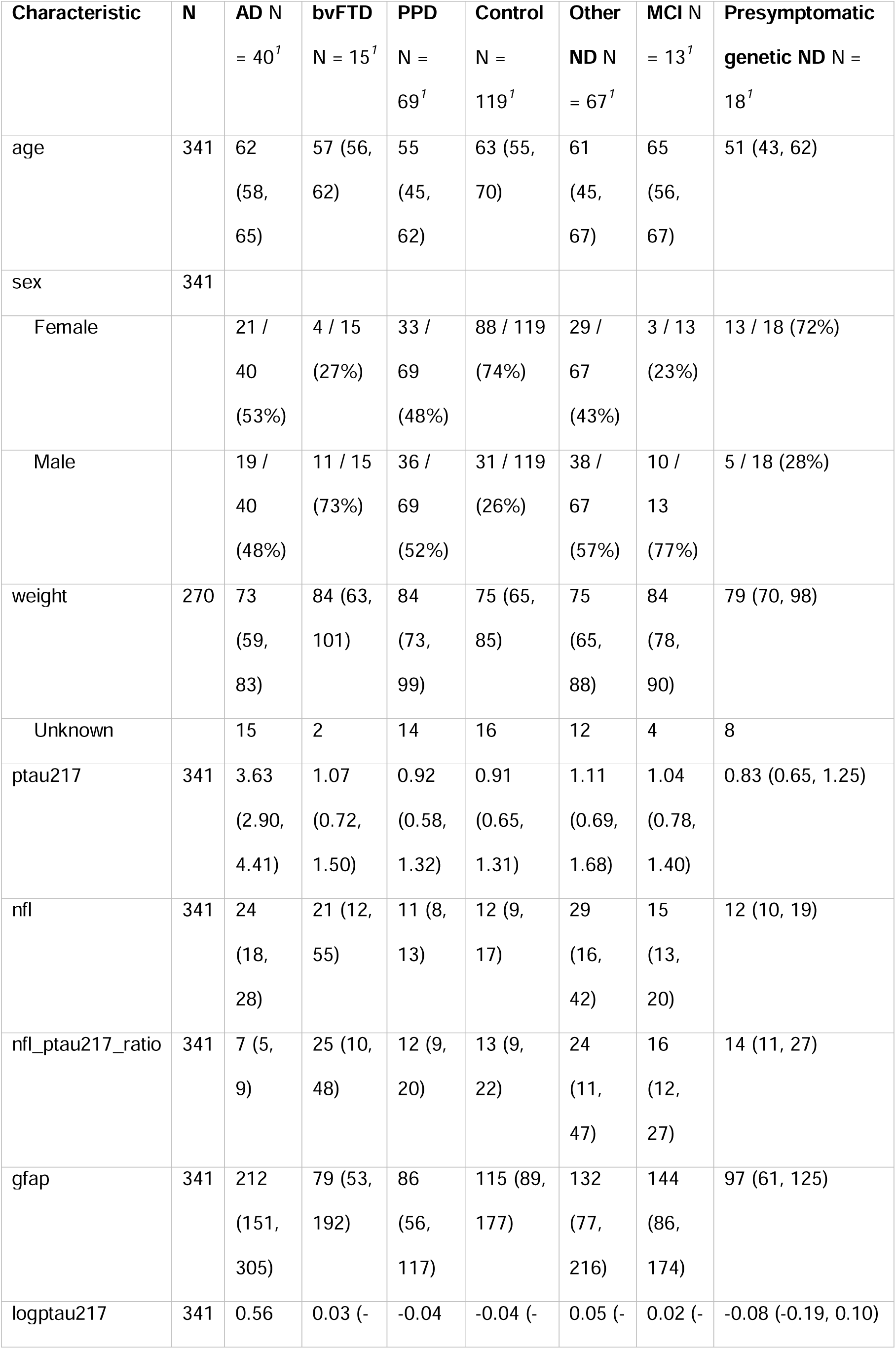
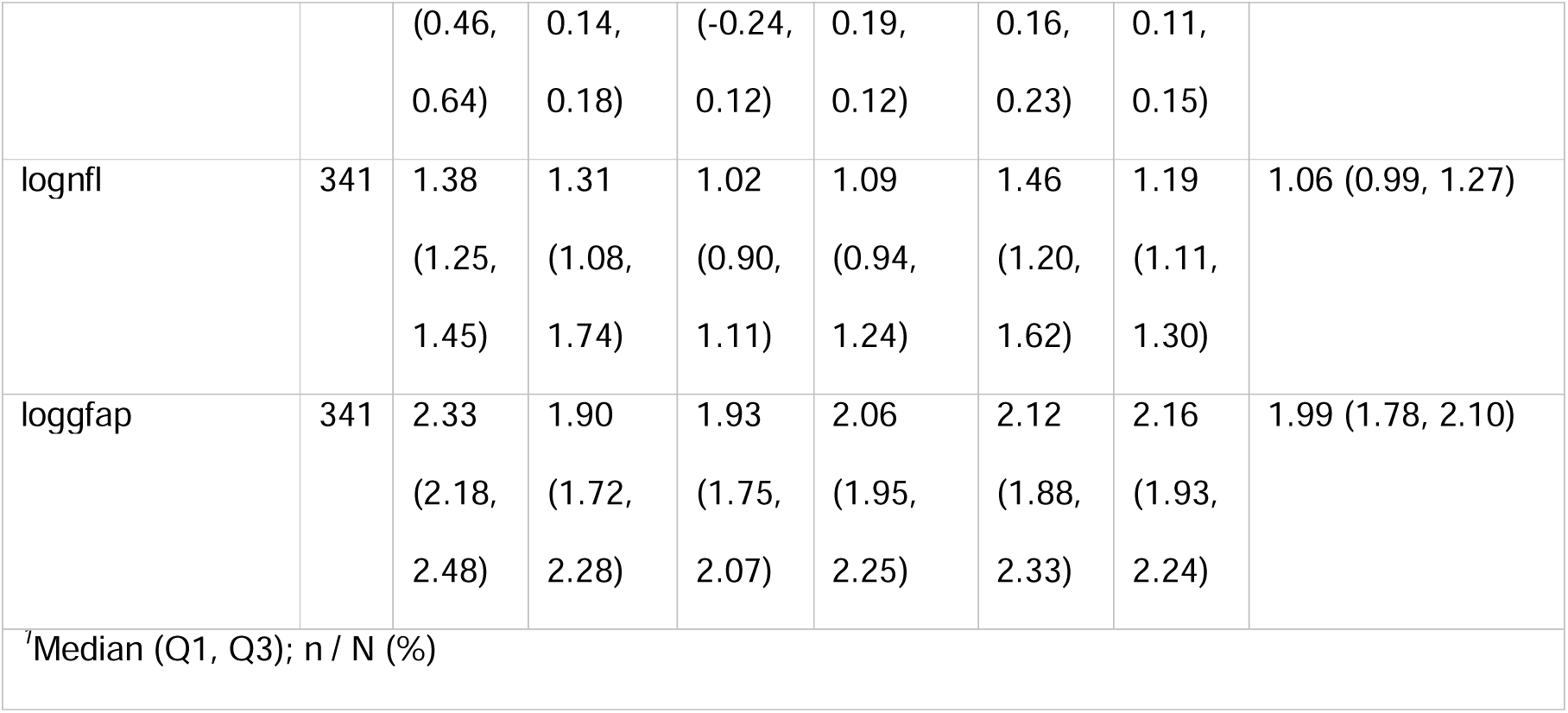
Study cohort details and biomarker levels.

PPD consisted of major depressive disorder (MDD, n=21), bipolar disorder (n=6), functional neurological/cognitive disorder (n=8), schizophrenia spectrum disorder (n=16), bvFTD ‘phenocopy’ syndrome (n=2), and other PPD (n=16, which included anxiety, personality, obsessive-compulsive, post-traumatic stress, and undifferentiated psychiatric disorders).

Other ND consisted of Creutzfeldt-Jakob disease (CJD, n=2), dementia with Lewy bodies (n=5), dementia not otherwise specified (n=6), Huntington disease (HD, n=15), mixed AD/vascular (n=3), substance-related cognitive impairment/dementia (n=2), vascular dementia (n=5), and a range of other ND (n=29, which included autoimmune encephalitis, cerebral amyloid angiopathy, corticobasal syndrome, CNS vasculitis, Down syndrome, Fahr disease, metabolic disorders, Niemann–Pick Type C, Parkinson’s disease, and cerebellar degenerative disorder). The presymptomatic genetic ND group consisted of: Alzheimer disease (PSEN1, n=2), genetic CJD (n=6), HD (n=6), CADASIL (n=1), bvFTD (C9orf72 n=1, GRN n=2).

80 patients had CSF biomarker analysis including AD proteins CSF AB42 and ptau181 (see Supplementary Material). Briefly, 27/40 (68%) of AD patients had CSF AD biomarkers. Most (18/27, 67%) had a CSF AD biomarkers profile consistent with Alzheimer disease, as defined by amyloid positive, phosphorylated tau positive (A+T+) profile, categorised using cut-offs as described in our previous study [10]. Other profiles in the AD group were 8/27 with A+T– and 1/27 with A-T+. 47% (7/15) of bvFTD had CSF AD biomarkers (none with an A+T+ profile, 4/7 with A-T-, 3/7 with A+T-). None of the patients in the other groups who had CSF AD biomarkers (20 in the PPD group, 21 in Other ND, 5 MCI) had A+T+ profiles (further details in Supplementary Material).

### Levels of plasma ptau217, NfL, NfL/ptau217 ratio, GFAP, in AD, bvFTD, and PPD

#### Plasma ptau 217

As demonstrated in Table 1 and Figure 1, plasma ptau217 levels were significantly elevated in AD compared to bvFTD (β=1.30 95%CI:[0.71, 1.73], p<0.001), and PPD (β=1.59 [1.34, 1.80], p<0.001). Levels were also elevated in AD compared to the other groups (Controls, Other ND, MCI, and Presymptomatic ND, all p<0.001).

**Figure 1.**
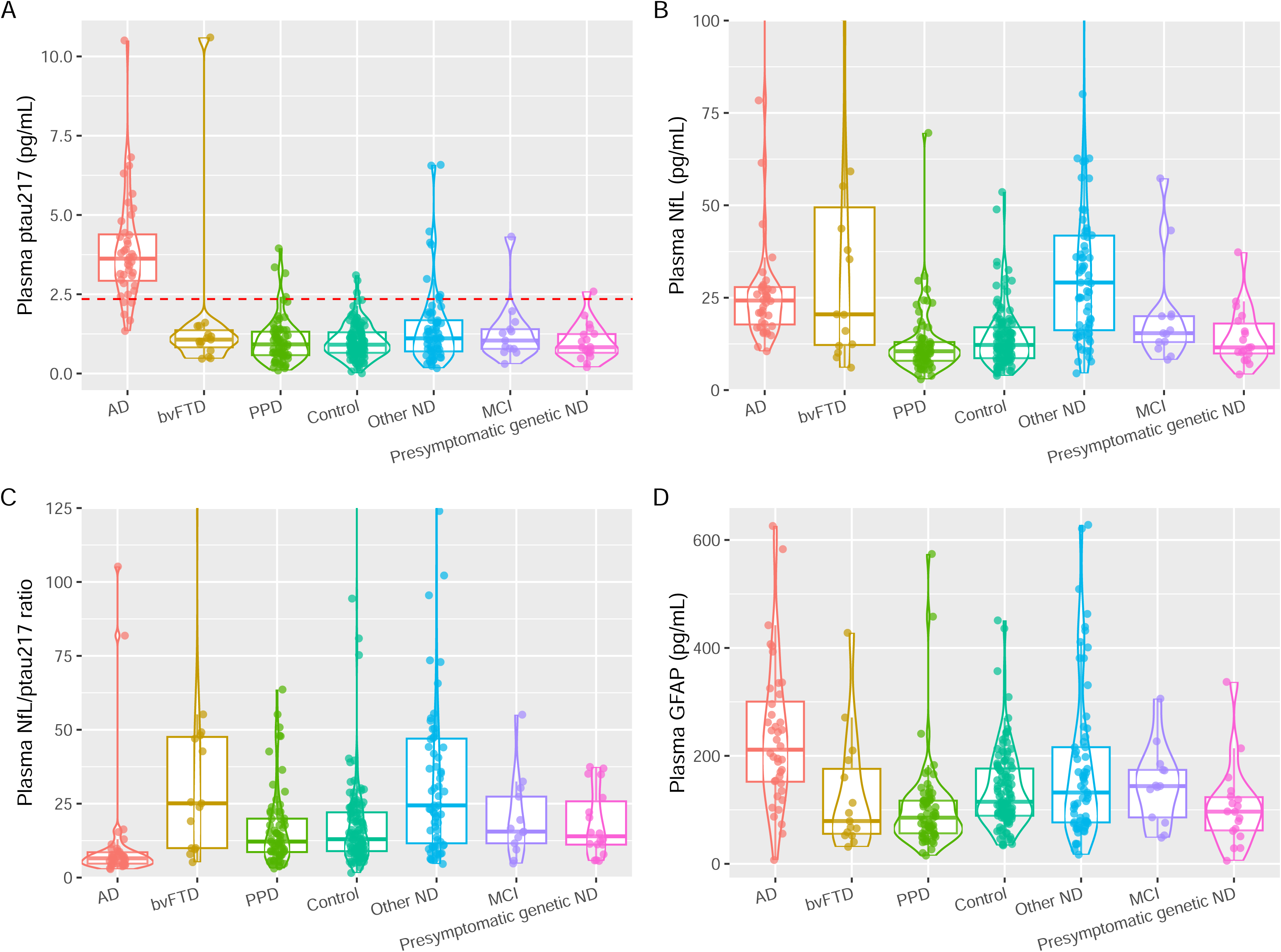
Plasma ptau217, NfL, NfL/ptau217 ratio, and GFAP levels, in AD, bvFTD, PPD, Controls, and other comparator groups. Plasma ptau217 levels were significant elevated in AD compared to all other disorders and controls. To improve readability, 7 outliers were not displayed in plot B for NfL (2 for AD (141 and 294pg/mL), 2 bvFTD (101 and 214pg/mL), 3 Other ND (106, 352, 1154pg/mL)), and 6 in plot C for NfL/ptau217 ratio (1 bvFTD (143), 2 controls (699, 772), 3 other ND (190, 198, 604). Dashed red lines = pre-defined optimal cut-offs from our previous studies, for ptau217 (2.35pg/mL).

#### Plasma NfL

Plasma NfL was elevated in AD compared to PPD (bootstrapped GLM with age and sex as additional covariates β=0.87 [0.59, 1.13], p<0.001), but not for AD compared to bvFTD (β=-0.10 [-0.82, 0.58], p=0.788). There were no significant differences in plasma NfL levels across the ND groups (AD, bvFTD, Other ND, all p>0.05). Levels were also similar between PPD, Control, MCI, and Presymptomatic ND groups.

#### The plasma NfL/ptau217 ratio

The plasma NfL/ptau217 ratio was reduced in AD, compared to bvFTD (β=-1.57 [-2.16, –0.94], p<0.001), and PPD (β=-0.99 [-0.58, –1.34], p<0.001). The ratio was also lower in AD compared to all the other groups (Controls, Other ND, MCI, Presymptomatic ND, all p<0.002).

#### Plasma GFAP

GFAP levels were elevated in AD compared to bvFTD (β=0.80 [0.23, 1.39], p=0.010), and PPD (β=0.82 [0.36, 1.21], p<0.001). GFAP levels was also elevated in AD compared to some other groups (Controls, MCI, Presymptomatic ND, all p<0.038), but not between AD and Other ND (p=0.108).

### Biomarker diagnostic performance to distinguish AD from PPD, AD from bvFTD, and bvFTD from PPD

#### Distinguishing AD from PPD

Ptau217 demonstrated the strongest diagnostic performance to distinguish AD from PPD, as demonstrated in Figure 2 and Supplementary Material. Ptau217 had an area under the curve (AUC) of 0.97, outperforming NfL, NfL/ptau217 ratio, and GFAP (AUCs 0.89, 0.77, 0.86 respectively; AUC differences all p<0.016). Ptau217 had the highest accuracy (93%), specificity (91%) and sensitivity (95%), at an optimal cutoff of 1.84pg/mL. Further details of all diagnostic performance metrics are available in Supplementary Material.

**Figure 2.**
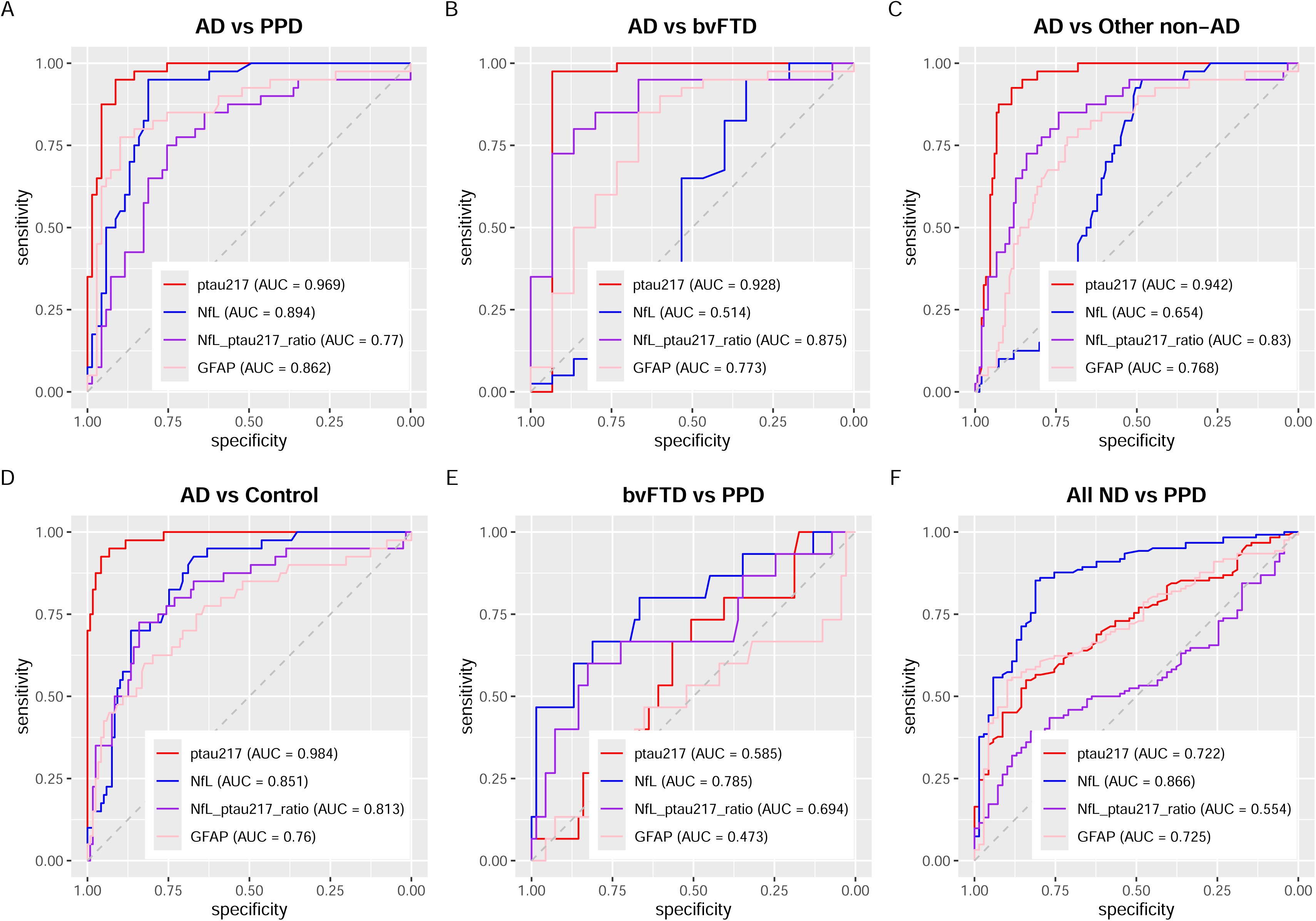
ROC analyses for diagnostic performance of plasma ptau217, NfL, NfL/ptau217 ratio, and GFAP. Plasma ptau217 had the very strong diagnostic performance for AD vs PPD, AD vs bvFTD, AD vs non-AD, AD vs controls, outperforming other biomarkers and the NfL/ptau217 ratio. NfL’s diagnostic performance was for bvFTD vs PPD, and All ND vs PPD.

#### Distinguishing AD from bvFTD

To distinguish AD from bvFTD, ptau217 once again demonstrated the highest AUC (0.93), and strongest diagnostic performance (93% specificity, 98% sensitivity, 96% accuracy, cut-off 1.64), compared to the other biomarkers. NfL did not have significant diagnostic performance to distinguish AD from bvFTD (AUC 0.51 [0.30, 0.74]). The AUC difference was not statistically different between ptau217 and the NfL/ptau217 ratio (AUC 0.88, AUC difference p=0.509), or GFAP (AUC 0.77, AUC difference p=0.10). However, compared to ptau217, both the ratio and GFAP demonstrated poorer specificity (87% and 67%, respectively), sensitivity (80% and 85%), and accuracy (82% and 80%), and much lower diagnostic odds ratios (26 and 11.33, versus ptau217’s 546).

#### Distinguishing AD from non-AD disorders

Ptau217 demonstrated high diagnostic performance to distinguish AD from other non-AD disorders (consisting of bvFTD, PPD, Other ND), with an AUC of 0.94. It was superior to NfL, NfL/ptau217 (all p less than or equal to 0.001), and had 88% specificity, 93% sensitivity, 90% accuracy, cut-off 2.19. Ptau217 demonstrated very strong diagnostic performance to distinguish AD from Controls (AUC 0.98, 96% specificity, 93% sensitivity).

#### Distinguishing bvFTD from PPD

For distinguishing bvFTD from PPD, NfL had the highest AUC (0.78), and strongest diagnostic performance (81% specificity, 67% sensitivity, 79% accuracy, cut-off 15.15pg/mL), although the AUC difference between NfL and NfL/ptau217 ratio was not significant (p=0.14). ptau217 and GFAP had no diagnostic utility (AUC confidence intervals both crossed 0.50).

### Biomarker diagnostic performance to distinguish all ND from PPD

As a follow on from our previous studies that focused on NfL in distinguishing ND from PPD, we investigated the ability of ptau217 and GFAP to distinguish ND as a group from PPD. We created an “All ND” group by combining AD, bvFTD, and Other ND, and compared this All ND group to PPD.

NfL demonstrated the highest AUC (0.87), and strongest diagnostic performance (81% specificity, 85% sensitivity, 84% accuracy, cut-off 14.35), significantly outperforming GFAP (AUC 0.73, AUC difference p<0.0001) and ptau217 (AUC 0.72, AUC difference p<0.001). The NfL/ptau217 ratio did not have diagnostic utility (AUC confidence interval crossed 0.50).

### Biomarker profiles in diagnostic groups

#### AD pathology/neuronal injury (Ptau217/NfL) biomarker profiles

We described ptau217 and NfL biomarker profiles in the diagnostic groups, using our previously described cutoff for plasma ptau217, and age-adjusted percentiles model for NfL [10,27].

As demonstrated in Figure 3, the high ptau217 profiles were most commonly seen in AD (total 88% of AD, 60% high ptau217/high NfL and 28% high ptau217/low NfL biomarker profiles). By comparison less than 10% of each of the other groups exhibited high ptau217 profile bvFTD only had 7% with high ptau217 profiles (all high ptau217/high NfL), PPD only 6% (3% high ptau217/high NfL, 3% high ptau217/low NfL), Controls only 3% (all high ptau217/low NfL), MCI and presympomatic ND 8% and 6% respectively. Other ND had 13% with high ptau217 profiles (10% high ptau217/high NfL, 3% high ptau217/low NfL).

**Figure 3.**
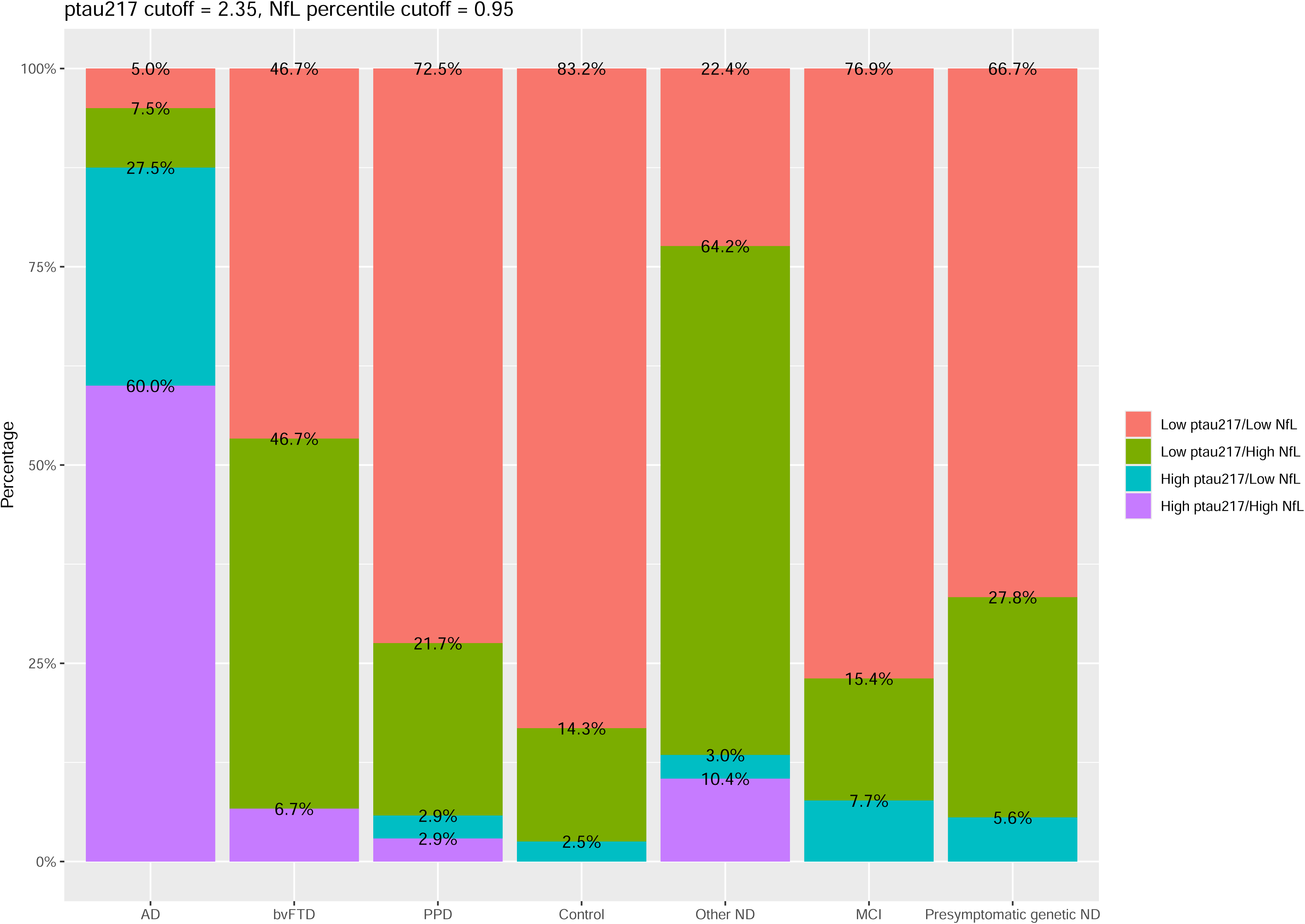
Ptau217 and NfL biomarker profiles, classified based on previously described cut-offs, in different diagnostic groups.

68% of AD had high NfL profiles (high ptau217/high NfL, low ptau217/high NfL), and AD had the lowest proportion of “normal” (low ptau217/low NfL) profiles. For bvFTD on the other hand, only 53% had high NfL profiles, and the remaining 47% all had low ptau217/low NfL profiles. Most PPD had a normal profile (72%), with 25% showing high NfL profiles. 83% of controls had a normal profile, and only 14% having high NfL profiles. Other ND had the highest proportions of high NfL levels (74%, 64% low ptau217/high NfL, 10% high ptau217/high NfL). Similar to PPD and controls, most participants in MCI, and presymptomatic ND had low ptau217/low NfL profiles (72%, 77%, 67% respectively).

These findings are further demonstrated in Figure 4, a plot of log ptau217 versus NfL age-adjusted z-score, as well as distributions. The scatter plot and distributions demonstrate that almost all AD patients had elevated ptau217 levels and high ptau217/high NfL profiles (upper right in scatter plot), as compared to non-AD disorders where almost all had low ptau217 levels. Most ND had high NfL levels, compared to a small proportion of PPD and Controls.

**Figure 4.**
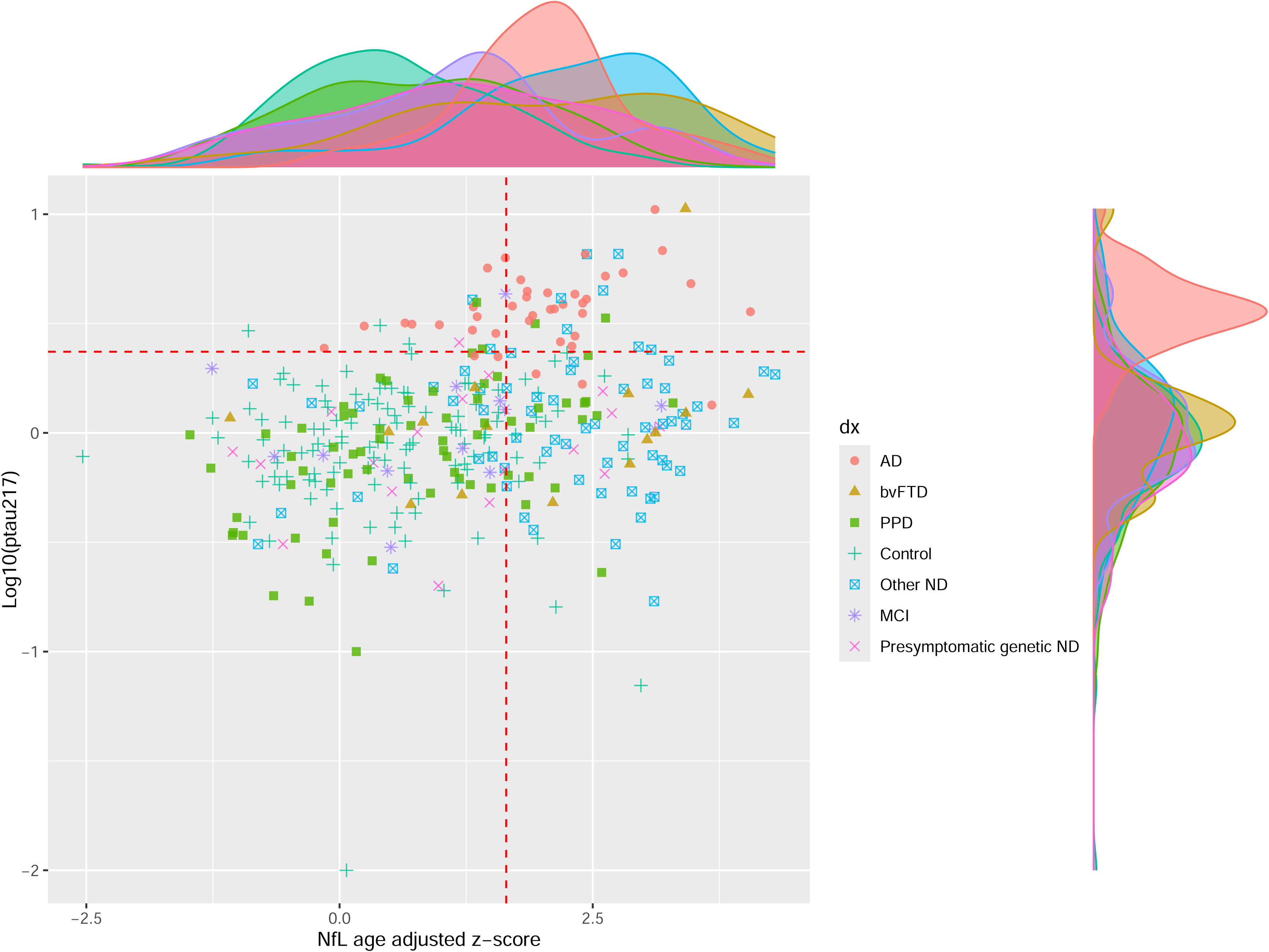
ptau217 and NfL levels and distributions in study cohort. The scatter plot and distributions demonstrate that almost all AD patients had elevated ptau217 levels and high ptau217/high NfL profiles, and compared to non-AD disorders where almost all had low ptau217 levels. Most ND had high NfL levels, compared to a small proportion of PPD and Controls. Dashed red lines = pre-defined optimal cut-offs from our previous studies, for ptau217 (log10 of 2.35pg/mL) and age-adjusted NfL (z-score of 1.645 or 95^th^ percentile).

#### AD pathology/neuronal injury/neuroinflammation (ptau217/NfL/GFAP) biomarker profiles

As an exploratory aim, we developed a novel GAMLSS age-based model for plasma GFAP to derive percentiles and z-scores, based on our control group. As demonstrated in Figure 5, this shows a similar non-linear association with age, and a u-shaped pattern similar to other studies [35,36]. This enabled complex Ptau217/NfL/GFAP (AD pathology/neuronal injury/neuroinflammation) biomarker profiling, which is demonstrated in Figure 6. Inclusion of GFAP to create this three biomarker profile, resulted in some differences compared to ptau217/NfL only profiling. The proportion of “normal” profiles using three biomarkers (i.e. Low ptau217/ Low NfL / Low GFAP) was lower in bvFTD (40%, compared to 47% Low ptau217 / Low NfL profiles). The proportion of normal ptau217/NfL/GFAP profiles in PPD, Controls, were similar to normal ptau217/NfL profiles.

**Figure 5.**
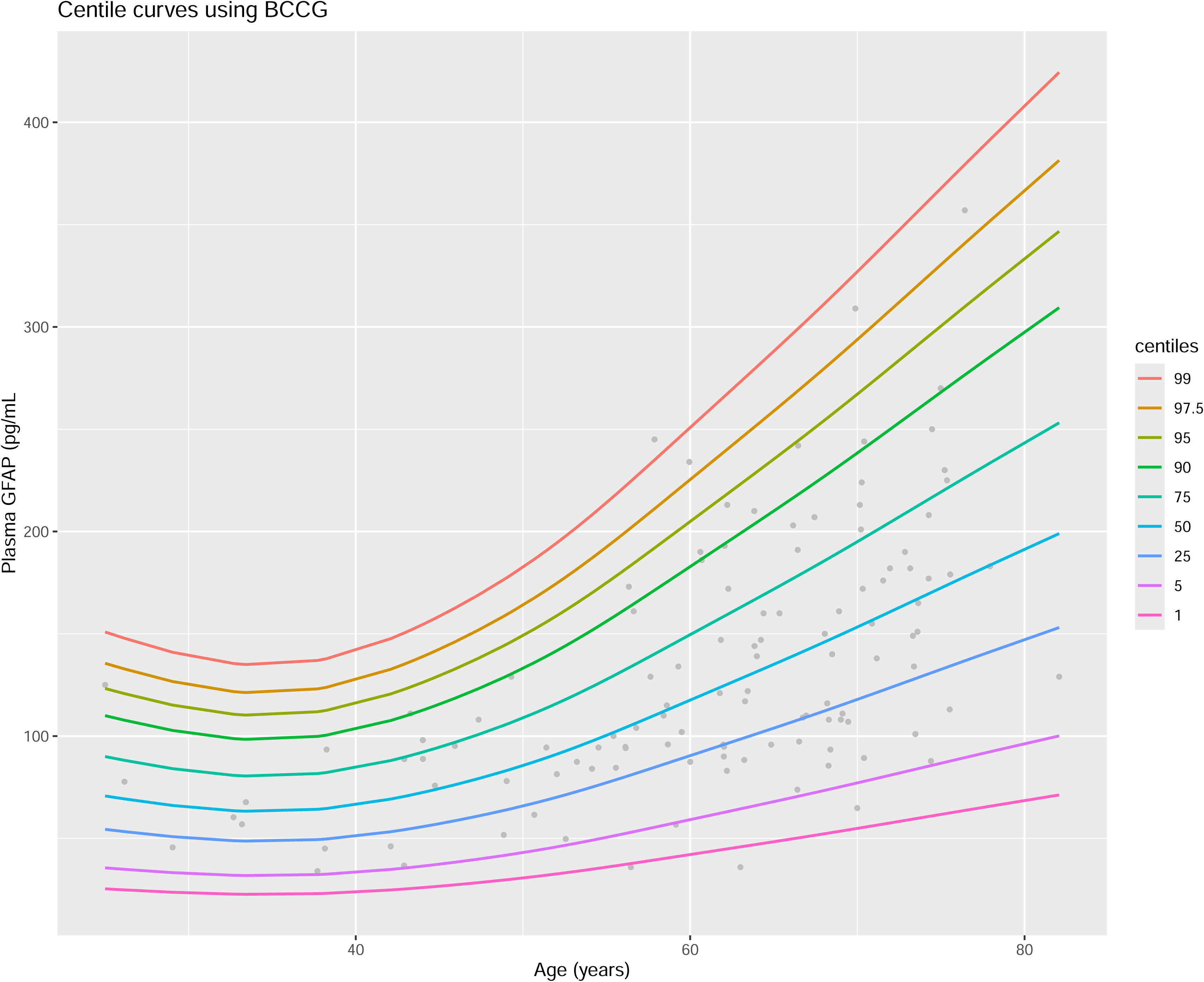
Plasma GFAP percentiles derived from generalised additive models for location, scale, and shape, based on this study’s control group. Two extreme outliers were excluded for modelling (GFAP levels > 400 pg/mL, aged 52 and 63), given the disproportionate influence they would have with the relatively small sample size.

**Figure 6.**
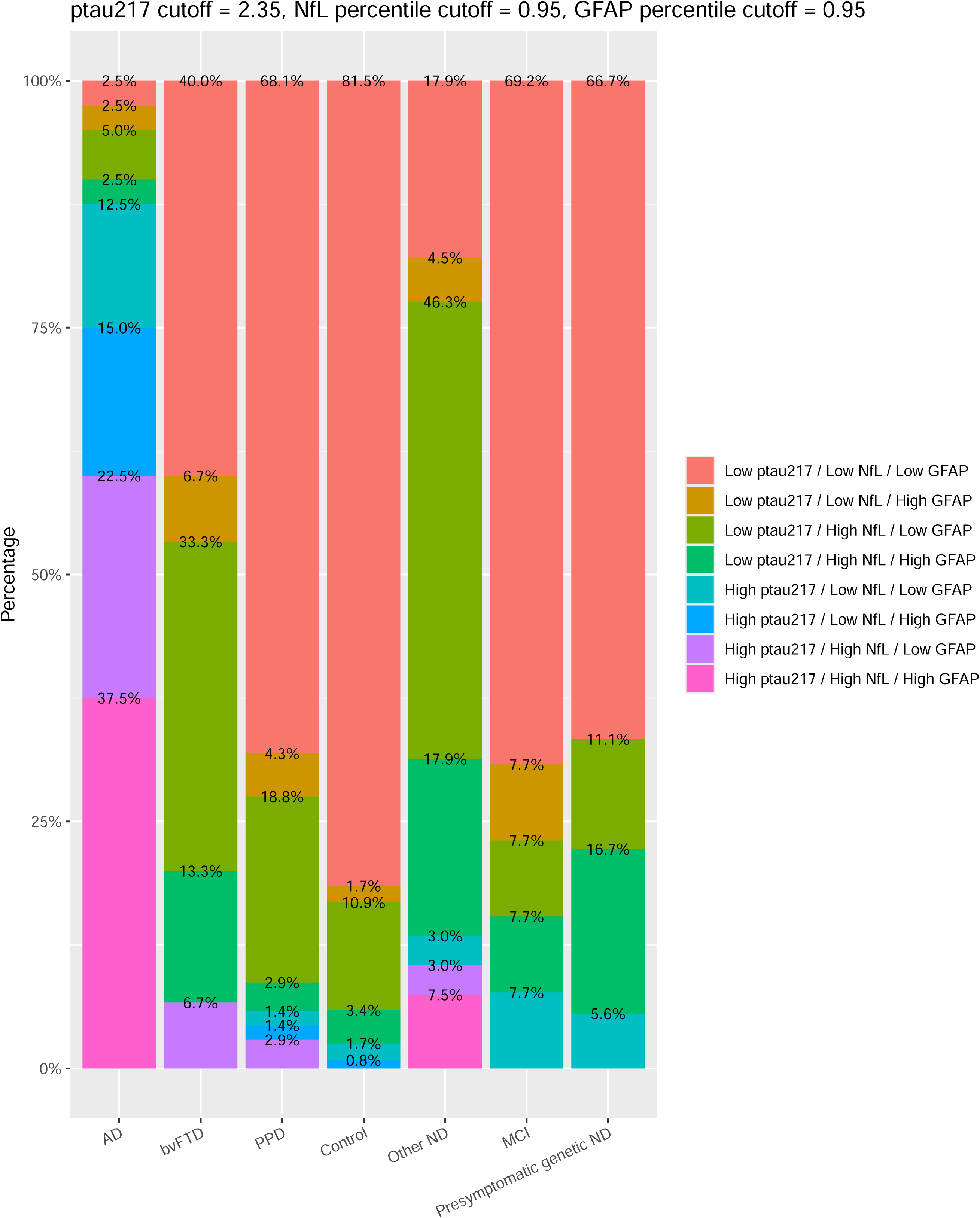
Ptau217, NfL and GFAP biomarker profiles in diagnostic groups.

## DISCUSSION

This study investigated multiple biomarkers in an unselected real-world clinical cohort of younger participants seen in a neuropsychiatry memory clinic and, focused on AD, bvTD, and PPD, with Controls, Other ND, MCI, and presymptomatic genetic ND as comparator groups. The main finding from this study was the very strong diagnostic performance of plasma ptau217 to distinguish AD from bvFTD, and AD from PPD. Second, ptau217 had superior performance for AD diagnosis on its own, compared to NfL/ptau217 ratio, NfL, and GFAP. Third, we found NfL to have the strongest performance for distinguishing bvFTD from PPD, and All ND from PPD. The findings of this study add important further evidence to the limited literature so far, of roles for different biomarkers in a real-world clinical cohort, with ptau217 being a very sensitive and specific test for AD, and NfL having strongest diagnostic performance to distinguish neurodegeneration from PPD/non-neurodegenerative disorders, with little role for GFAP in any of these distinctions. The key strengths of this study included a real-world, unselected diverse, clinical and younger cohort, which are a relatively under-investigated group. Establishing the role for these biomarkers in younger people, who face greater rates of misdiagnosis and diagnostic uncertainty, is important for increasing clinical and research access to this group where arguably the clinical and broader psychosocial benefits could be greater.

This study builds on our previous work on ptau217 [10], and provides further evidence of the very high specificity of plasma ptau217 for AD, and very strong diagnostic performance for some of the most common diagnostic distinctions, distinguishing AD from bvFTD, and AD from PPD. Notably, we saw very high specificity and sensitivity (91% and 95%), establishing the role for such an accurate test (93%) in younger people. While we hypothesised that an NfL/ptau217 ratio could improve distinction between AD and bvFTD, and bvFTD from PPD, we found no benefit of NfL/ptau217 ratio. This finding contrasts to that of Benussi et al, who found some benefit of NfL/ptau217 ratio compared to ptau217 alone [31]. This difference was slight, and may reflect greater FTD sample sizes, and older ages, in that cohort. While there has been some interest in GFAP assisting with AD vs non-AD and ND vs PPD/non-ND distinctions, we found little to no utility for GFAP.

An additional strength of our, study was using cut-offs and reference ranges that we previously published [10,27,38], to define and describe biomarker profiles in different diagnostic groups. Although the previous ptau217 study and cutoff had some overlap with this study cohort, it is important to note that a new separate analysis of AD vs controls in this study cohort, yielded a very similar cutoff of (2.18pg/mL, Supplementary Material). The NfL reference range was from a completely independent large reference cohort [27,38]. Rousset et al also described diagnostic performance of ptau217 and ptau217/NfL biomarker profiles. We found higher diagnostic performance for ptau217 (AUC 0.97 in our study for AD vs PPD, and 0.98 for AD vs controls, sensitivities and specificities and accuracies >90%), compared to their study (AUCs approximately 0.78-0.81 depending on the group being compared to AD). We also found greater proportions of high ptau217/high NfL and high ptau217/low NfL profiles in our AD group. Our findings may reflect our slightly younger AD group (median 62 years of age, versus 66). While most patients with PPD had low ptau217/low NfL profiles, there were still a number with other profiles, including 25% with high NfL. This builds on numerous other studies that have demonstrated, that while very elevated NfL levels are rarely seen in PPD, slightly elevated levels can be seen in numerous PPDs and PPDs cannot be assumed to be equivalent to controls/non-neurological controls [3,4,25–27,30,39].

In the ND groups (comprising AD, bvFTD, Other ND), we found that bvFTD had the greatest proportion (47%) of “normal” ptau217/NfL biomarker profiles (Low ptau217 / Low NfL). This was reduced slightly when considering “normal” ptau217/NfL/GFAP (Low ptau217/ Low NfL / Low GFAP) profile (40%). While this could reflect technical limitations and issues previously described with technical/analytical issues and reference ranges issues, it most likely also highlights the greater complexity and clinical and biomarker heterogeneity, seen in bvFTD compared other disorders, and the ongoing challenges faced in this clinical diagnosis [40]. While ptau217 can accurately distinguish frontal AD from bvFTD, and NfL has strong diagnostic performance for bvFTD vs PPD at group levels, many individuals with bvFTD would still have non-diagnostic biomarker levels and profiles. Studies are needed and underway to more deeply understand this discordance in larger groups of people with bvFTD, and by including multimodal neuroimaging, clinical, multi-omic and genomic data, to ultimately improve upon single diagnostic biomarker performance for more accurate diagnosis of bvFTD. It is important to note that describing biomarker profiles were secondary, exploratory aims of this study, and a significant limitation is the binary/dichotomous classification. Problems with dichotomous classification important limitations we and others have identified regarding cut-offs and reference ranges [14,41], losing important nuance in interpreting an individual level – for example and individual with level 1% below the binary cut-off or 95^th^ percentile, would be classified as “low”, the same as someone with a level 50% below the cutoff or 95^th^ percentile. Unfortunately, the small sample sizes prevented more nuanced categorisation in this study.

There was a very high ptau217 outlier level in the bvFTD group. This patient had a complex history. They were initially diagnosed with Alzheimer disease based on memory impairment and clinical and neuroimaging features. The patient subsequently developed significant personality and behaviour change, and eventually developed motor neurone disease. At this point, the clinical diagnosis was revised to bvFTD with MND. Notably, their blood sample and biomarker level was prior to the onset of MND. It is possible that their elevated ptau217 level reflected a skeletal muscle source of ptau217 [42], or it could suggest that they was unfortunate to have multiple pathologies, such as AD, as well as MND, and possibly an FTD as well. Unfortunately, this patient did not have genetic testing, or CSF AD protein analysis or amyloid PET. It is possible that blood-based biomarkers such as ptau217 could help not only improve accurate AD diagnosis, but also improve identification of cases of multiple co-pathologies. These issues are especially important for older people, where co-pathology is more common, and in this era of increasingly available disease specific treatments [43].

Limitations of this study include the single-centre and cross-sectional nature of the study, the relatively small FTD subgroup sample, the lack of gold standard reference tests for AD (CSF AD proteins and amyloid PET) for all participants, and the lack of definitive genetic or post-mortem diagnostic confirmation. The focus of this study was on AD, bvFTD, and PPD groups. While Other ND, MCI, and presymptomatic genetic ND were included as comparator groups for secondary/exploratory/descriptive analyses, their sample sizes limited more any further in-depth analyses or interpretations. A novel aspect of our study was developing a GAMLSS model for plasma GFAP. We found a ‘u-shape’ curve (higher levels in younger people, lower in midlife, then increasing again with age) similar to other studies [35,36], however, our model based on a relatively small number of individuals, and therefore should be interpreted with caution. Future studies are underway, aiming replicate these findings and to investigate the incremental validity of multiple biomarkers in a larger sample size, and in different clinical settings (in particular, in low prevalence settings such as primary psychiatric and primary care settings).

This study adds important evidence for the very high specificity, sensitivity, and overall diagnostic performance of ptau217 to distinguish AD from bvFTD and PPD. In addition, we further establish the role of plasma NfL, and superiority over other biomarkers, for broadly distinguishing ND from PPD/non-neurodegenerative disorders, in younger patients in clinical settings,. This study’s findings, of the strength and simplicity of single blood-based biomarker tests for AD diagnosis is particularly important in the era of increasing availability of anti-amyloid and other disease specific treatments for AD. Ongoing further research will further establish the role of ptau217 and NfL for precision diagnostic algorithms to reduce misdiagnosis and diagnostic delay, and improve clinical and research outcomes.

## Data Availability

All data produced in the present study are available upon reasonable request to the authors

## Keywords

## ACKNOWLEDGEMENTS AND FUNDING SOURCES

The authors would like to thank all the patients, participants, and their families for their participation.

KB is supported by the Swedish Research Council (#2017-00915 and #2022-00732), the Swedish Alzheimer Foundation (#AF-930351, #AF-939721, #AF-968270, and #AF-994551), Hjärnfonden, Sweden (#ALZ2022-0006, #FO2024-0048-TK-130 and FO2024-0048-HK-24), the Swedish state under the agreement between the Swedish government and the County Councils, the ALF-agreement (#ALFGBG-965240 and #ALFGBG-1006418), the European Union Joint Program for Neurodegenerative Disorders (JPND2019-466-236), the Alzheimer’s Association 2021 Zenith Award (ZEN-21-848495), the Alzheimer’s Association 2022-2025 Grant (SG-23-1038904 QC), La Fondation Recherche Alzheimer (FRA), Paris, France, the Kirsten and Freddy Johansen Foundation, Copenhagen, Denmark, Familjen Rönströms Stiftelse, Stockholm, Sweden, and an anonymous filantropist and donor.

HZ is a Wallenberg Scholar and a Distinguished Professor at the Swedish Research Council supported by grants from the Swedish Research Council (#2023-00356, #2022-01018 and #2019-02397), the European Union’s Horizon Europe research and innovation programme under grant agreement No 101053962, Swedish State Support for Clinical Research (#ALFGBG-71320), the Alzheimer Drug Discovery Foundation (ADDF), USA (#201809-2016862), the AD Strategic Fund and the Alzheimer’s Association (#ADSF-21-831376-C, #ADSF-21-831381-C, #ADSF-21-831377-C, and #ADSF-24-1284328-C), the European Partnership on Metrology, co-financed from the European Union’s Horizon Europe Research and Innovation Programme and by the Participating States (NEuroBioStand, #22HLT07), the Bluefield Project, Cure Alzheimer’s Fund, the Olav Thon Foundation, the Erling-Persson Family Foundation, Familjen Rönströms Stiftelse, Familjen Beiglers Stiftelse, Stiftelsen för Gamla Tjänarinnor, Hjärnfonden, Sweden (#FO2022-0270), the European Union’s Horizon 2020 research and innovation programme under the Marie Skłodowska-Curie grant agreement No 860197 (MIRIADE), the European Union Joint Programme – Neurodegenerative Disease Research (JPND2021-00694), the National Institute for Health and Care Research University College London Hospitals Biomedical Research Centre, the UK Dementia Research Institute at UCL (UKDRI-1003), and an anonymous donor.

The corresponding author had full access to all the data in the study and had final responsibility for the decision to submit for publication.

A.F.S is primarily funded by the Swedish federal government under the ALF agreement (ALF 2022 YF 0017) and The Åke Wiberg Foundation

