## Supplementary Material for "Plasma ptau217, NfL, GFAP diagnostic performance and biomarker profiles in Alzheimer disease, frontotemporal dementia, and psychiatric disorders, in a prospective unselected neuropsychiatry memory clinic"

| Characteristic | N | **AD** N = 40[1] | **bvFTD** N = 15[1] | **PPD** N = 69[1] | **Control** N = 119[1] | **Other ND** N = 67[1] | **MCI** N = 13[1] | **Presymptomatic genetic ND** N = 18[1] |
|---|---|---|---|---|---|---|---|---|
| age | 341 | 62 (58, 65) | 57 (56, 62) | 55 (45, 62) | 63 (55, 70) | 61 (45, 67) | 65 (56, 67) | 51 (43, 62) |
| sex | 341 | | | | | | | |
| Female | | 21 / 40 (53%) | 4 / 15 (27%) | 33 / 69 (48%) | 88 / 119 (74%) | 29 / 67 (43%) | 3 / 13 (23%) | 13 / 18 (72%) |
| Male | | 19 / 40 (48%) | 11 / 15 (73%) | 36 / 69 (52%) | 31 / 119 (26%) | 38 / 67 (57%) | 10 / 13 (77%) | 5 / 18 (28%) |
| weight | 270 | 73 (59, 83) | 84 (63, 101) | 84 (73, 99) | 75 (65, 85) | 75 (65, 88) | 84 (78, 90) | 79 (70, 98) |
| Unknown | | 15 | 2 | 14 | 16 | 12 | 4 | 8 |
| ptau217 | 341 | 3.63 (2.90, 4.41) | 1.07 (0.72, 1.50) | 0.92 (0.58, 1.32) | 0.91 (0.65, 1.31) | 1.11 (0.69, 1.68) | 1.04 (0.78, 1.40) | 0.83 (0.65, 1.25) |
| nfl | 341 | 24 (18, 28) | 21 (12, 55) | 11 (8, 13) | 12 (9, 17) | 29 (16, 42) | 15 (13, 20) | 12 (10, 19) |
| nfl_ptau217_ratio | 341 | 7 (5, 9) | 25 (10, 48) | 12 (9, 20) | 13 (9, 22) | 24 (11, 47) | 16 (12, 27) | 14 (11, 27) |
| gfap | 341 | 212 (151, 305) | 79 (53, 192) | 86 (56, 117) | 115 (89, 177) | 132 (77, 216) | 144 (86, 174) | 97 (61, 125) |
| logptau217 | 341 | 0.56 (0.46, 0.64) | 0.03 (-0.14, 0.18) | -0.04 (-0.24, 0.12) | -0.04 (-0.19, 0.12) | 0.05 (-0.16, 0.23) | 0.02 (-0.11, 0.15) | -0.08 (-0.19, 0.10) |

| Characteristic | N | **AD** N = 40[1] | **bvFTD** N = 15[1] | **PPD** N = 69[1] | **Control** N = 119[1] | **Other ND** N = 67[1] | **MCI** N = 13[1] | **Presymptomatic genetic ND** N = 18[1] |
|---|---|---|---|---|---|---|---|---|
| lognfl | 341 | 1.38 (1.25, 1.45) | 1.31 (1.08, 1.74) | 1.02 (0.90, 1.11) | 1.09 (0.94, 1.24) | 1.46 (1.20, 1.62) | 1.19 (1.11, 1.30) | 1.06 (0.99, 1.27) |
| loggfap | 341 | 2.33 (2.18, 2.48) | 1.90 (1.72, 2.28) | 1.93 (1.75, 2.07) | 2.06 (1.95, 2.25) | 2.12 (1.88, 2.33) | 2.16 (1.93, 2.24) | 1.99 (1.78, 2.10) |
| csf_status | 80 | | | | | | | |
| A-T- | | 0 / 27 (0%) | 4 / 7 (57%) | 12 / 20 (60%) | 0 / 0 (NA%) | 6 / 21 (29%) | 1 / 5 (20%) | 0 / 0 (NA%) |
| A-T+ | | 1 / 27 (3.7%) | 0 / 7 (0%) | 1 / 20 (5.0%) | 0 / 0 (NA%) | 2 / 21 (9.5%) | 2 / 5 (40%) | 0 / 0 (NA%) |
| A+T- | | 8 / 27 (30%) | 3 / 7 (43%) | 7 / 20 (35%) | 0 / 0 (NA%) | 13 / 21 (62%) | 2 / 5 (40%) | 0 / 0 (NA%) |
| A+T+ | | 18 / 27 (67%) | 0 / 7 (0%) | 0 / 20 (0%) | 0 / 0 (NA%) | 0 / 21 (0%) | 0 / 5 (0%) | 0 / 0 (NA%) |
| Unknown | | 13 | 8 | 49 | 119 | 46 | 8 | 18 |
| amyloid_status | 80 | | | | | | | |
| A- | | 1 / 27 (3.7%) | 4 / 7 (57%) | 13 / 20 (65%) | 0 / 0 (NA%) | 8 / 21 (38%) | 3 / 5 (60%) | 0 / 0 (NA%) |
| A+ | | 26 / 27 (96%) | 3 / 7 (43%) | 7 / 20 (35%) | 0 / 0 (NA%) | 13 / 21 (62%) | 2 / 5 (40%) | 0 / 0 (NA%) |
| Unknown | | 13 | 8 | 49 | 119 | 46 | 8 | 18 |
| ad_status | 80 | | | | | | | |

| **Characteristic** | **N** | **AD** N = 40[1] | **bvFTD** N = 15[1] | **PPD** N = 69[1] | **Control** N = 119[1] | **Other ND** N = 67[1] | **MCI** N = 13[1] | **Presymptomatic genetic ND** N = 18[1] |
|---|---|---|---|---|---|---|---|---|
| A+T+ | | 18 / 27 (67%) | 0 / 7 (0%) | 0 / 20 (0%) | 0 / 0 (NA%) | 0 / 21 (0%) | 0 / 5 (0%) | 0 / 0 (NA%) |
| Other | | 9 / 27 (33%) | 7 / 7 (100%) | 20 / 20 (100%) | 0 / 0 (NA%) | 21 / 21 (100%) | 5 / 5 (100%) | 0 / 0 (NA%) |
| Unknown | | 13 | 8 | 49 | 119 | 46 | 8 | 18 |

[1]Median (Q1, Q3); n / N (%)

| Categorisation | Difference | AUC | Cutoff | Spec | Sens | LR+ | LR- | PPV | NPV | DOR | Accuracy |
|---|---|---|---|---|---|---|---|---|---|---|---|
| **AD vs PPD** | | | | | | | | | | | |
| ptau217 | >NfL (p=0.016) >ratio (p<0.0001) >GFAP (p=0.010) | 0.97 [0.94, 1.00] | 1.84 | 91% | 95% | 10.93 | 0.05 | 86% | 97% | 199.5 | 93% |
| NfL | <ptau217 | 0.89 [0.83, 0.95] | 14.55 | 81% | 95% | 5.04 | 0.06 | 75% | 75% | 81.8 | 86% |

| Categorisation | Difference | AUC | Cutoff | Spec | Sens | LR+ | LR- | PPV | NPV | DOR | Accuracy |
|---|---|---|---|---|---|---|---|---|---|---|---|
| NfL/ptau217 ratio | <ptau217 | 0.77 [0.68, 0.87] | 8.57 | 75% | 75% | 3.04 | 0.33 | 84% | 84% | 9.18 | 75% |
| GFAP | <ptau217 | 0.86 [0.78, 0.94] | 148.5 | 90% | 78% | 7.64 | 0.25 | 82% | 87% | 30.51 | 85% |
| **AD vs bvFTD** | | | | | | | | | | | |
| ptau217 | >NfL (p<0.001) | 0.93 [0.80, 1.00] | 1.64 | 93% | 98% | 14.63 | 0.03 | 98% | 93% | 546 | 96% |
| NfL | | 0.51 [0.30, 0.74] | | | | | | | | | |
| NfL/ptau217 ratio | | 0.88 [0.77, 0.98] | 9.27 | 87% | 80% | 6 | 0.23 | 94% | 92% | 26 | 82% |
| GFAP | | 0.77 [0.62, 0.93] | 115.5 | 67% | 85% | 2.55 | 0.23 | 87% | 62% | 11.33 | 80% |
| **AD vs Other non-AD** | | | | | | | | | | | |
| ptau217 | >NfL (p<0.001) >ratio (p=0.001) | 0.94 [0.91, 0.98] | 2.19 | 88% | 93% | 8.21 | 0.08 | 69% | 98% | 97.21 | 90% |

| Categorisation | Difference | AUC | Cutoff | Spec | Sens | LR+ | LR- | PPV | NPV | DOR | Accuracy |
|---|---|---|---|---|---|---|---|---|---|---|---|
| | >GFAP (p<0.001) | | | | | | | | | | |
| NfL | | 0.65 [0.58, 0.73] | 14.6 | 48% | 95% | 1.84 | 0.10 | 33% | 97% | 17.78 | 58% |
| NfL/ptau217 ratio | >NfL (p=0.009) | 0.83 [0.75, 0.91] | 9.87 | 74% | 85% | 3.29 | 0.20 | 47% | 95% | 16.27 | 76% |
| GFAP | >NfL (p=0.017) | 0.77 [0.69, 0.85] | 148.5 | 72% | 78% | 2.72 | 0.31 | 42% | 92% | 8.65 | 73% |
| **bvFTD vs PPD** | | | | | | | | | | | |
| ptau217 | | 0.59 [0.43, 0.74] | | | | | | | | | |
| NfL | | 0.78 [0.64, 0.93] | 15.15 | 81% | 67% | 3.54 | 0.41 | 43% | 92% | 8.62 | 79% |
| NfL/ptau217 ratio | | 0.69 [0.52, 0.86] | 23.13 | 83% | 60% | 3.45 | 0.48 | 43% | 90% | 7.13 | 79% |
| GFAP | | 0.43 [0.29, 0.66] | | | | | | | | | |

| Categorisation | Difference | AUC | Cutoff | Spec | Sens | LR+ | LR- | PPV | NPV | DOR | Accuracy |
|---|---|---|---|---|---|---|---|---|---|---|---|
| **AD+bvFTD vs PPD** | | | | | | | | | | | |
| ptau217 | >ratio (p<0.0001) >GFAP (p=0.026) | 0.86 [0.80, 0.93] | 1.47 | 84% | 78% | 4.90 | 0.26 | 80% | 83% | 18.89 | 81% |
| NfL | >ratio (p=0.001) >GFAP (p=0.044) | 0.86 [0.80, 0 93] | 14.55 | 81% | 87% | 4.63 | 0.16 | 79% | 89% | 29.54 | 84% |
| NfL/ptau217 ratio | <ptau217 <NfL | 0.64 [0.54, 0.75] | 8.57 | 75% | 58% | 2.36 | 0.55 | 65% | 69% | 4.26 | 68% |
| GFAP | <ptau217 <NfL | 0.77 [0.68, 0.86] | 148.5 | 90% | 65% | 6.45 | 0.38 | 84% | 77% | 16.78 | 79% |
| **ND vs PPD** | | | | | | | | | | | |
| ptau217 | <NfL | 0.72 [0.65, 0.79] | 1.45 | 84% | 55% | 3.44 | 0.54 | 86% | 51% | 6.42 | 65% |

| Categorisation | Difference | AUC | Cutoff | Spec | Sens | LR+ | LR- | PPV | NPV | DOR | Accuracy |
|---|---|---|---|---|---|---|---|---|---|---|---|
| NfL | >ptau217 (p<0.001) >GFAP (p<0.0001) | 0.87 [0.81, 0.92] | 14.35 | 81% | 85% | 4.52 | 0.18 | 89% | 86% | 24.89 | 84% |
| NfL/ptau217 ratio | | 0.55 [0.47, 0.64] | | | | | | | | | |
| GFAP | <NfL | 0.73 [0.65, 0.80] | 148.5 | 90% | 55% | 5.41 | 0.50 | 91% | 53% | 10.79 | 68% |
| **Additional comparisons** | | | | | | | | | | | |
| **AD vs Controls** | | | | | | | | | | | |
| ptau217 | | 0.98 [0.97, 1.00] | 2.18 | 96% | 93% | 22.02 | 0.08 | 88% | 97% | 281.2 | 95% |
| NfL | | 0.85 [0.79, 0.91] | 15.25 | 87% | 93% | 2.82 | 0.11 | 49% | 96% | 25.30 | 74% |
| NfL/ptau217 ratio | | 0.81 [0.73, 0.90] | 7.81 | 84% | 73% | 4.54 | 0.33 | 60% | 90% | 13.88 | 81% |

| Categorisation | Difference | AUC | Cutoff | Spec | Sens | LR+ | LR- | PPV | NPV | DOR | Accuracy |
|---|---|---|---|---|---|---|---|---|---|---|---|
| GFAP | | 0.76 [0.67, 0.85] | 192 | 82% | 60% | 3.4 | 0.49 | 53% | 86% | 7 | 77% |
| **All ND vs Controls** | | | | | | | | | | | |
| ptau217 | | 0.72 [0.65, 0.78] | 1.84 | 93% | 45% | 6.71 | 0.59 | 87% | 62% | 11.39 | 69% |
| NfL | | 0.82 [0.77, 0.87] | 20.15<br>14.7 | 87%<br>63% | 66%<br>84% | 4.88<br>2.28 | 0.40<br>0.25 | 83%<br>70% | 71%<br>80% | 12.26<br>9.24 | 76%<br>74% |
| NfL/ptau217 ratio | | 0.52 [0.45, 0.60] | | | | | | | | | |
| GFAP | | 0.59 [0.52, 0.66] | 191.5 | 82% | 41% | 2.32 | 0.72 | 70% | 58% | 3.24 | 61% |
| **AD+bvFTD vs Controls** | | | | | | | | | | | |
| ptau217 | | 0.87 [0.80, 0.94] | 2.18 | 96% | 69% | 16.44 | 0.32 | 88% | 87% | 50.96 | 87% |
| NfL | | 0.82 [0.75, 0.89] | 20.15 | 87% | 67% | 5.00 | 0.38 | 70% | 85% | 13.23 | 80% |
| NfL/ptau217 ratio | | 0.68 [0.58, 0.78] | 7.96 | 83% | 56% | 3.35 | 0.52 | 61% | 80% | 6.39 | 75% |

| Categorisation | Difference | AUC | Cutoff | Spec | Sens | LR+ | LR- | PPV | NPV | DOR | Accuracy |
|---|---|---|---|---|---|---|---|---|---|---|---|
| GFAP | | 0.66 [0.56, 0.76] | 191.5 | 82% | 51% | 2.88 | 0.60 | 57% | 78% | 4.84 | 72% |
